# Total body weight estimation by 3D camera systems: potential high-tech solutions for emergency medicine applications? A scoping review

**DOI:** 10.1101/2024.08.14.24311987

**Authors:** Mike Wells, Lara Nicole Goldstein, Terran Wells, Niloufar Ghazi, Abhijit Pandya, Borifoje Furht, Gabriella Engstrom, Muhammad Tanveer Jan, Richard Shih

## Abstract

**Background:** Weight estimation is required in adult patients when weight-based medication must be administered during emergency care, as measuring weight is often not possible. Inaccurate estimations may lead to inaccurate drug dosing, which may cause patient harm. High-tech 3D camera systems driven by artificial intelligence might be the solution to this problem. The aim of this review was to describe and evaluate the published literature on 3D camera weight estimation methods.

**Methods:** A systematic literature search was performed for articles that studied the use of 3D camera systems for weight estimation in adults. Data on the study characteristics, the quality of the studies, the 3D camera methods evaluated, and the accuracy of the systems were extracted and evaluated.

**Results:** A total of 14 studies were included, published from 2012 to 2024. Most studies used Microsoft Kinect cameras, with various analytical approaches to weight estimation. The 3D camera systems often achieved a P10 of 90% (90% of estimates within 10% of actual weight), with all systems exceeding a P10 of 78%. The studies highlighted a significant potential for 3D camera systems to be suitable for use in emergency care.

**Conclusion:** The 3D camera systems offer a promising method for weight estimation in emergency settings, potentially improving drug dosing accuracy and patient safety. Weight estimates were extremely accurate. Importantly, 3D camera systems possess characteristics that could make them very appropriate for use during emergency care. Future research should focus on developing and validating this methodology in larger studies with true external and clinical validation.

## INTRODUCTION

### Background

During the resuscitative care of critically ill or injured patients, an estimation of their weight is required when weight-based drug therapy is required, and actual patient weight cannot be measured. Measuring weight with scales is not always feasible as it is time-consuming and requires patient cooperation as well as a medically stable patient [1]. If a stand-on scale cannot be used due to the patient’s clinical condition, some emergency departments use scales that are imbedded in patient stretchers. These are costly, not universally available, and unproven in terms of accuracy during emergency care [2, 3]. An estimation of weight is, therefore, often required.

Numerous studies have described or evaluated different methods of weight estimation in adults. These methods include (1) estimates by patients themselves, by family members, and by healthcare professionals; (2) formulas based on anthropometric measurements, such as the Lorenz formula; (3) dual length- and habitus-based tapes, such as the PAWPER XL-MAC tape; and (4) high-tech methods, such as 3D camera systems. Of all these methods, 3D camera systems have shown the greatest potential for highly accurate, rapid, easy-to-use estimation of weight [4]. These methods use real-time 3D images and previously trained artificial intelligence algorithms to generate estimates of weight. Existing 3D camera weight estimation methods have used different approaches with different cameras, software, costs, ease-of-use, applicability, and accuracy.

### Importance

Inaccurate weight estimations may lead to inaccurate drug doses. This may cause patient harm through ineffective treatment (underestimation of weight) or adverse drug effects (overestimation of weight) [5]. One weight estimation researcher has written: “It cannot be considered to be good medical practice to use a weight estimation system that is known to be inaccurate” [6]. In adults, most current methods of weight estimation are simply not accurate enough [4]. It is possible that 3D camera methods of weight estimation may offer a solution to these problems. In addition, simple and easy-to-use methods are of special interest in emergency medicine. This is because more complex methods may be less easy to use and more prone to errors during the high cognitive loads experienced in emergency care [7]. High-tech 3D camera systems engineered for usability in the setting of emergency care may also be able to address this problem.

### Goals of this investigation

Our aim in this scoping review was to review the available literature in which a 3D camera system was used to estimate a patient’s weight, with a medical indication as the ultimate purpose. We aimed to describe the performance and accuracy of 3D camera weight estimation systems, the types of cameras used, and the analytical and software methods used in the weight estimation process.

## METHODS

This scoping review was based on the PRISMA for Scoping Reviews guidelines (PRISMA-ScR) [8].

### Literature search

A literature search was conducted using MEDLINE, EMBASE, IEEE Xplore, and Google Scholar. Eligible studies published between January 2012 and April 2024 were identified using the search strategy shown in Supplementary Table 1.

### Eligibility criteria

Studies were included for further evaluation if they were peer reviewed, full length, English language papers containing original data. Studies evaluating any form of 3D camera weight estimation methodology, and in any type of participants were eligible for inclusion if an accurate measured weight was used as the standard reference. Studies on weight estimation not relevant to a clinical or hospital setting were excluded (e.g., weight estimation for forensic or non-medical applications).

### Selection of studies

The titles and abstracts of the articles identified by the database search were manually screened by two researchers independently (MW, NG). The full texts of the selected reviews were then obtained and assessed for eligibility. Any differences in opinion were resolved by discussion and consensus.

### Critical appraisal of individual sources of evidence (Grading of quality of studies)

Every included study was graded for quality of evidence using a modified Newcastle-Ottawa scale (NOS), as has been described previously (see Supplementary Table 2) [9]. Studies were downgraded if significant methodological weaknesses were present, e.g., if data presentation was incomplete or if performance outcome data was not appropriately presented or analyzed. An assessment of selective non-reporting or under-reporting of results in the studies was included in the Newcastle-Ottawa scale. Each study could score a minimum of zero stars and a maximum of 10 stars on the modified NOS. On this scale, a study with score from 6 to 10 has high quality, 4 to 5 has a moderate risk of bias, and 0 to 3 a very high risk of bias.

### Data charting process (Data extraction)

Data extraction was conducted by one researcher (TW) using a standardized electronic data extraction form and was independently confirmed by another researcher (MW) for accuracy.

### Data items

The following data was extracted: basic study information (region of origin, study population, sample size), study participant characteristics, 3D camera used, analytic method or software used for the weight estimation process, key findings, and the data presented on the performance or accuracy of weight estimation.

### Data synthesis (Map of outcomes)

The findings of this scoping review were synthesized by presenting a descriptive and quantitative summary of the study characteristics using frequencies with percentages. The studies were grouped by the types of 3D cameras used, as well as the overall analytic approach, and summarized according to weight estimation outcomes.

In terms of the quantitative analysis, the main outcomes of interest were metrics representing the performance of the weight estimation system. These included mean error or mean percentage error, which represented the estimation bias; the root mean square error, the mean absolute error, the root mean square percentage error or the mean absolute percentage error, which quantified the estimation precision; and the percentage of weight estimations that fell within 10% (P10) as well as within 20% (P20) of measured weight, which denoted overall accuracy. We considered the measures of overall accuracy (P10 and P20) to be the best indicator of overall performance, as we have described previously [10]. If P10 data was not reported it was imputed, whenever possible, from other reported metrics (mean absolute percentage error or mean percentage error).

## RESULTS

No significant deviations from the protocol were noted. The details of the numbers of sources of evidence screened, assessed for eligibility, and included in the review, with reasons for exclusions at each stage are shown in Figure 1.

**Figure 1.**
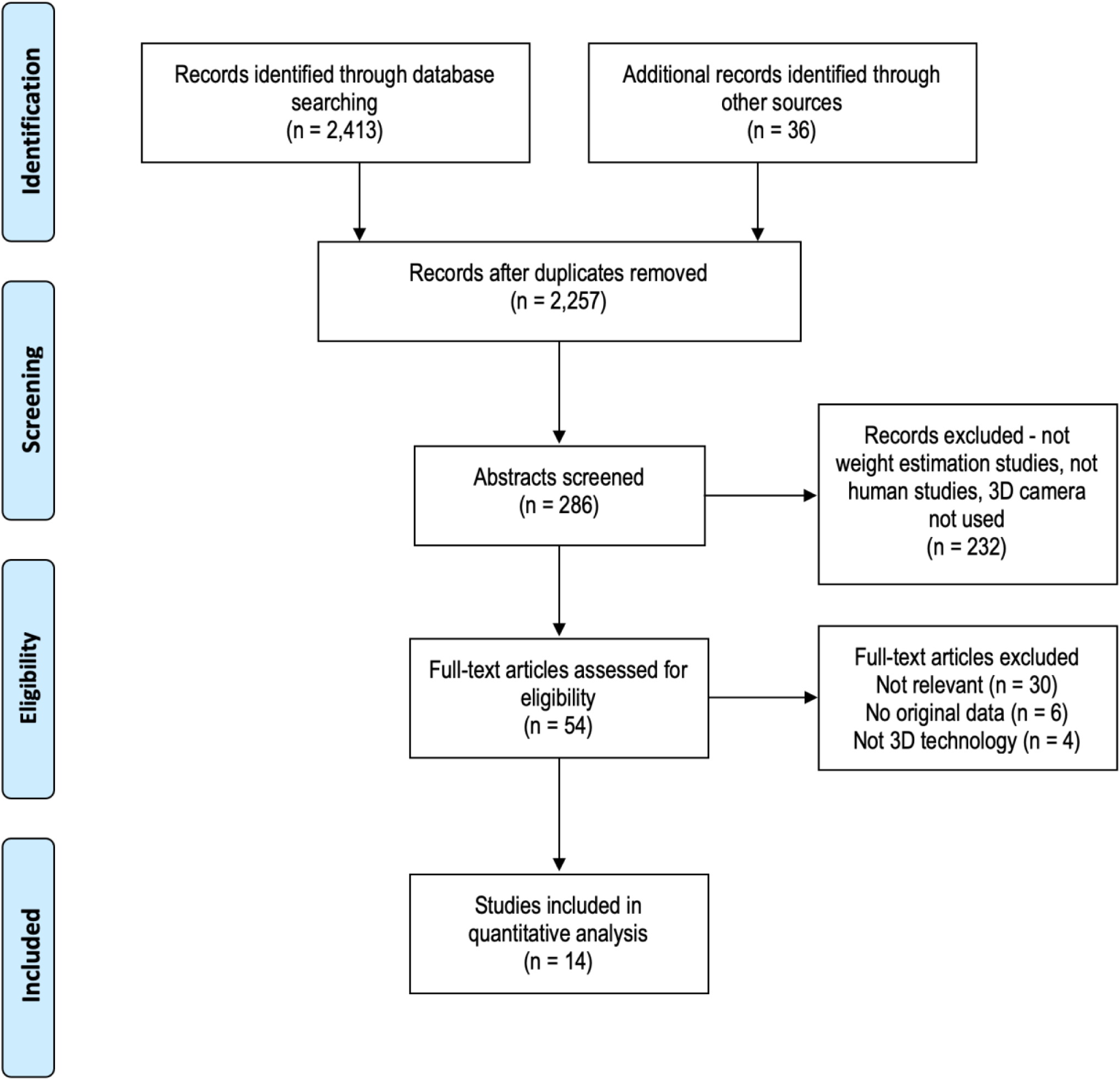
The Preferred Reporting Items for Scoping Reviews (PRISMA-ScR) flow chart for article identification and selection.

### Characteristics of the included studies

A total of 14 studies were included in this scoping review. The details of the included studies, including the study methodologies, the hardware and software used, and the weight estimation approaches are shown in Table 1. Two thirds of the studies (9/14 (64%)) were from Europe (all but one from Germany), with three studies (21%) from the USA, and two studies (14%) from elsewhere (one from Indonesia and one from Chile).

**Table 1.** Details of the included studies [11–24]. Abbreviations: NHANES – National Health and Nutrition Examination Survey; BMI – Body Mass Index; MATLAB-MATrix LABoratory; PCL – Point Cloud Library; ANN – Artificial Neural Network; MAPE – Mean Absolute Percentage Error; RANSAC – RANdom SAmple Consensus; SVR – Support Vector Regression; LibSVM – library for support vector machines; MAE – Mean Absolute Error; MPE – Mean Percentage Error; LOA – Limits of Agreement; P10 – percentage of estimates within 10% of actual weight; LMS – Least Mean Squares; CNN – Convoluted Neural Network; DBSCAN – Density-Based Spatial Clustering of Applications with Noise; ADAM – Adaptive Moment Estimation.

| Author, date, location | Study population | Participant characteristics | Camera technology | Method of weight estimation | Analytic method or software | Accuracy data | Notes |
| --- | --- | --- | --- | --- | --- | --- | --- |
| Velardo *et al* 2012<br>France | NHANES 1999 to 2005 datasets.<br>N=28,000<br>Volunteers for testing of actual 3D camera system<br>N=15. | Age: >20 years.<br>Sex: not reported.<br>Weight: not reported.<br>BMI: not reported. | Microsoft Kinect 1 | Biometrics obtained from 3D image silhouette – regression equation to predict weight. User input required: none. | MATLAB Neural network toolbox to obtain estimators. OpenNI framework, Primesense, OpenCV, PCL. ANN (single hidden layer) to create algorithm for weight estimation. | **Weight:**<br>MAPE 3.6% in 10/15 human test subjects, using 3D camera. No data reported for females. | Gender recognition was included using neural network classifier on anthropometric measurements (80% accurate). No results reported for other 5/15 subjects. Internal validation: none. |
| Nguyen *et al* 2014<br>USA | University student and staff volunteers. N=190 | Age: 18-60 years.<br>Sex: not reported.<br>Weight: not reported.<br>BMI: not reported. | Microsoft Kinect 1 | Feature fusion models – including linear dimension features, area features, and sideview shape from 3D camera. User input required: none. | RANSAC for video processing. SVR- LibSVM with Gaussian Radial Basis Function for weight estimation. | **Weight:**<br>MAE 4.6kg (F), 5.6kg (M). MAE 5.4kg when autodetection of gender was used. | Gender estimation – 88-92% accuracy. The 3D camera system was more accurate than observer estimates. Internal validation: 5-fold cross validation. |
| Pfützner *et al* 2015<br>Germany | Convenience sample of Emergency Department patients. N=110 | Age: 19-86 years.<br>Sex: male 53.6%.<br>Weight: 49-117kg.<br>BMI: 18-40kg/m<sup>2</sup>. | Microsoft Kinect 1 & Optris PI400 (thermal camera) | The approach involves patient segmentation, body volume estimation and body weight estimation using a fixed coefficient for body density. User input required: gender. | RANSAC, point cloud geometry. | **Weight:**<br>MPE 1.02 (95% LOA - 15.8 to 17.9)<br>P10 79.1% | The 3D camera method was more accurate than physician estimates and similar to the Lorenz method, but less accurate than patient self-estimates. Internal validation: none. |
| Pfützner *et al* 2016<br>Germany | Convenience sample of Emergency Department patients. N=69 | Age: 18-87 years.<br>Sex: not reported.<br>Weight: 49-129kg.<br>BMI: 19-48kg/m<sup>2</sup>. | Microsoft Kinect 1 & Optris PI400 (thermal camera) | The approach used for weight estimation involves sensor fusion of an RGB-D sensor and a thermal camera, pre-processing and segmentation of the sensor data, extraction of ten features for machine learning-based weight estimation. User input required: gender. | Multiple features extracted from cloud point data. ANN (single hidden layer) to create algorithm for weight estimation. ANN trained using a dataset recorded in real emergency scenarios. | **Weight:**<br>MPE -0.7 (95% LOA - 13.5 to 12.1)<br>P10 89.9% | The 3D camera method was more accurate than physician estimates and the Lorenz method, but less accurate than patient self-estimates. Internal validation: none. |
| Benalcázar *et al* 2017<br>Chile | Volunteers N=185 (LMS method)<br>Volunteers N=34 (ANN method) | LMS model:<br>Age: 3-23 years.<br>Sex: male 100%.<br>Weight: 11-114kg.<br>BMI: not reported.<br>ANN model:<br>Age: 3-18 years.<br>Sex: male 100%. | Microsoft Kinect 1 | The area of the person in the normalized image was computed, and this parameter was used to estimate the weight of the person. User input required: information on hairstyle and clothing. | Both LMS and ANN fitting techniques were explored for the weight estimation model. The Levenberg Marquardt back-propagation method was used for training. | **Weight:**<br>LMS MAPE 10.7%<br>ANN MAPE 5.8% | The increase in accuracy between LMS and ANN was due to the change in evaluation of hair and clothing. Internal validation: k-fold cross-validation. |
|  |  | Weight: 17-88kg.<br>BMI: not reported. |  |  |  |  |  |
| Pfützner *et al* 2017<br>Germany | Convenience sample of trauma room patients<br>N=127. Volunteers from public event<br>N=106. | Age: not reported.<br>Sex: male 41.2%.<br>Weight: 49-129kg.<br>BMI: not reported. | Microsoft Kinect 2 | The features used for weight estimation include volume, surface area, number of points, density, eigenvalues, sphericity, flatness, linearity, compactness, kurtosis, alternative compactness, distance to person, contour length, contour area, convex hull length, convex hull area, gender, and temperature features. User input required: gender. | RANSAC for video processing, Artificial Neural Network for weight estimation. | **Weight:**<br>MPE 0.3 (95% LOA - 10.1 to 10.7)<br>P10 94.8% | The Kinect 2 was more accurate than the Kinect original and was perhaps underappreciated in the paper.<br>Internal validation: none. |
| Pfützner *et al* 2018<br>Germany | Various sources<br>N=299 (reanalysis of previous depth data with new methodology) | Age: not reported.<br>Sex: male 67.5%.<br>Weight: 49-129kg.<br>BMI: not reported. | Microsoft Kinect 1, Microsoft Kinect 2, Optris PI400) | New approach was used in which deep learning was used for the weight estimation process. Features are extracted from the person's point cloud, including geometric features, features based on eigenvalues, statistical features, and features from the silhouette of a person. These features are then used as input to different algorithms, such as clustering, a three-layer feedforward neural network, and an ANN, to estimate the body weight. User input required: gender. | CNN, ANN, RANSAC | **Weight:**<br>P10 95.3% for lying subjects, P10 91.3% for walking subjects and P10 100% for walking subjects. | This paper is closer to being a summary of other tests than being a standalone paper, although new modelling is used. The only new data is for walking subjects. Internal validation: none. |
| Bigalke *et al* 2021<br>Germany | Not reported.<br>N=60 for training, N=49 for testing | Age: not reported.<br>Sex: not reported.<br>Weight: 44-105kg.<br>BMI: not reported. | Not reported* | The approach used for weight estimation in this study was deep learning techniques applied to 3D point cloud data without relying on hand-crafted features. They adopt the concept of basis point sets (BPS) to encode the input point cloud into a low-dimensional feature vector, which is then passed to a neural network trained for weight regression. User input required: none. | RANSAC for image isolation, DBSCAN, ADAM optimizer, PointNet, fully connected neural network | **Weight:**<br>MAE 4.2 (0.12) kg<br>MAPE 6.4 (0.2) %<br>P10 78.6% | Many "not reported" items. Internal validation: split sample. |
| Dane 2021 *et al*<br>USA | Convenience sample of outpatient CT scan patients N=363 for training, N=90 for testing | Age: 59.8 (14.9) years.<br>Sex: not reported.<br>Weight: 34-107kg.<br>BMI: not reported. | FAST 3D camera (Siemens Healthineers) | The 3D camera captured the patient's body surface landmarks using infrared imaging. The patient's body was divided into different regions (head, thorax, abdomen, arm, and leg) based on the 3D patient geometry. From the estimated 3D patient body mesh, various geometry-based features such as volume and length of each body region were computed. | Deformable Patient Avatar (digital twin) with Deep Image Network.<br>The weight estimation was modeled as a weighted sum of all the geometry-based features, and the weight coefficients were estimated using a Bayesian Ridge regression model. This | **Height:**<br>MAPE 2.0% (1.4)<br>**Weight:**<br>MAPE 5.1% (4.3)<br>9.2% underweight (n=7)<br>5.4% normal weight (n=57)<br>4.6% obese (n=22) | Poorer estimations in underweight patients. Incomplete accuracy data reporting. Internal validation: split sample. |
|  |  |  |  | These features were used for weight estimation. User input required: none. | process was performed using Scikit-learn. |  |  |
| Geissler *et al* 2021<br>Germany | Random patients undergoing CT scanning N=221 for training, N=101 for testing. | Age: 21 to 92 years.<br>Sex: not reported.<br>Weight: not reported.<br>BMI: 27.3 (SD 5.5) kg/m<sup>2</sup>. | Microsoft Kinect 2 | Digital twin or avatar fitted to observed depth data, sized according to height. User input required: none. | Not reported. | **Height:**<br>MAE 2.5 (1.9) cm<br>**Weight:**<br>MAE 4.4 (3.9) kg<br>35% of patients had estimate error >5kg | Worse estimates in obese patients, but height estimates not affected. Weight estimates much worse than patient self-estimates, but much better than staff estimates. Internal validation: split sample. |
| Mameli *et al* 2021<br>Italy | Volunteers.<br>N=94 for training, N=9 for testing. | Age: not reported.<br>Sex: male 63.1%.<br>Weight: 40-100kg.<br>BMI: not reported. | Orbec Astra S2 | "Top View Weight Estimation Approach", VRAI Weight estimation dataset. Deep learning model trained directly off 3D depth data. User input required: none. | Deep Neural Networks (VGG16, ResNet, Inception DenseNet, EfficientNet) | **Weight:**<br>MAE \| MSE<br>ResNet 4kg \| 36kg<br>Inception 3kg \| 11kg<br>DenseNet 1kg \| 4kg<br>EfficientNet 1kg \| 2kg | Top view (bird's eye view) of standing participants was the only 3D image used in this study. Internal validation: split sample. |
| Naufal *et al* 2021<br>Indonesia | Volunteers N=147. | Age: 5-70 years.<br>Sex: not reported.<br>Weight: 14-90kg.<br>BMI: not reported. | Microsoft Kinect 1 | 3D images segmented and converted to 2D images. This image area was correlated with weight. User input required: N/A. | MATLAB-based software | **Weight:**<br>Only correlation was used. No valid method of assessing accuracy. | Height estimation accuracy to within 1%<br>Internal validation: none. |
| Tamersoy *et al* 2023<br>Germany | Volunteers plus patients undergoing CT or MRI imaging N=1850. | Age: not reported.<br>Sex: not reported.<br>Weight: 45-120kg.<br>BMI: not reported. | 3D camera not specified | The method treats the estimation of patient height and weight as separate single-value regression problems, eliminating the need for error-prone intermediate stages such as volume computations. A 3D patient avatar or digital twin image is fitted to the acquired depth images, which is then used for part-volume based weight estimation. User input required: none. | ResNet 18<br>ADAM optimizer | **Height:**<br>P5 – 98.4%<br>P15 – 99.9%<br>**Weight:**<br>P10 – 95.6%<br>P20 – 99.8% | Extremely high weight estimation accuracy. May possibly be overfitted model. Internal validation: 23-fold cross validation. |
| Shahzadi *et al* 2024<br>Germany | Consecutive patients undergoing MRI N=148. | Age: not reported.<br>Sex: not reported.<br>Weight: 45-120kg.<br>BMI: not reported. | myExam™ 3D Camera<br>Siemens Healthineers | Details not reported. Unspecified features from depth data used in separate prediction models for height and weight. Model was trained on an unspecified dataset of images. User input required: none. | ResNet18 for initial training. SMAPE as a loss function and ADAM optimizer. | **Height:**<br>P5 100%<br>MAPE 1.7 (1.2)%<br>**Weight:**<br>P10 85.1%<br>P20 95.9%<br>MAPE 5.6 (5.5)% | Without blanket P10 93.8, P20 100%. Height estimations also slightly better. Predictions were still good in patients with Class 2 obesity. Internal validation: 23-fold cross validation. |

Most of the studies (8/14 (57%)) prospectively collected data for data analysis, while six (43%) studies used existing data to develop or evaluate new analytic approaches. Only 4/14 (29%) studies compared 3D camera methods against other methods of weight estimation. The main aim of the study was to evaluate potential methods to estimate weight for drug dosing purposes in 6/14 (43%) studies, for CT contrast and radiation dosing in 5/14 (36%) studies, for nutritional or body habitus assessment in 2/14 (14%) studies, and other reasons in 1/14 (7%) study.

### Risks of bias and limitations across studies

The methodological quality of most of the studies was good. Most studies (11/14 (79%)) had a low risk of bias on the Newcastle-Ottawa scale, one study (7%) had moderate risk of bias and two studies (14%) had a high risk of bias. However, incomplete data reporting or incomplete statistical analysis (5/14 studies (36%)) were common. Only 3/14 (21%) studies presented any form of subgroup analyses and no study provided comprehensive subgroup analyses by sex and weight-status. In addition, 2/14 (14%) studies had a sample size of 100 or fewer participants, and only 3/14 (21%) studies had a sample size of greater than 300 participants. These findings are summarized in Figure 2.

**Figure 2.**
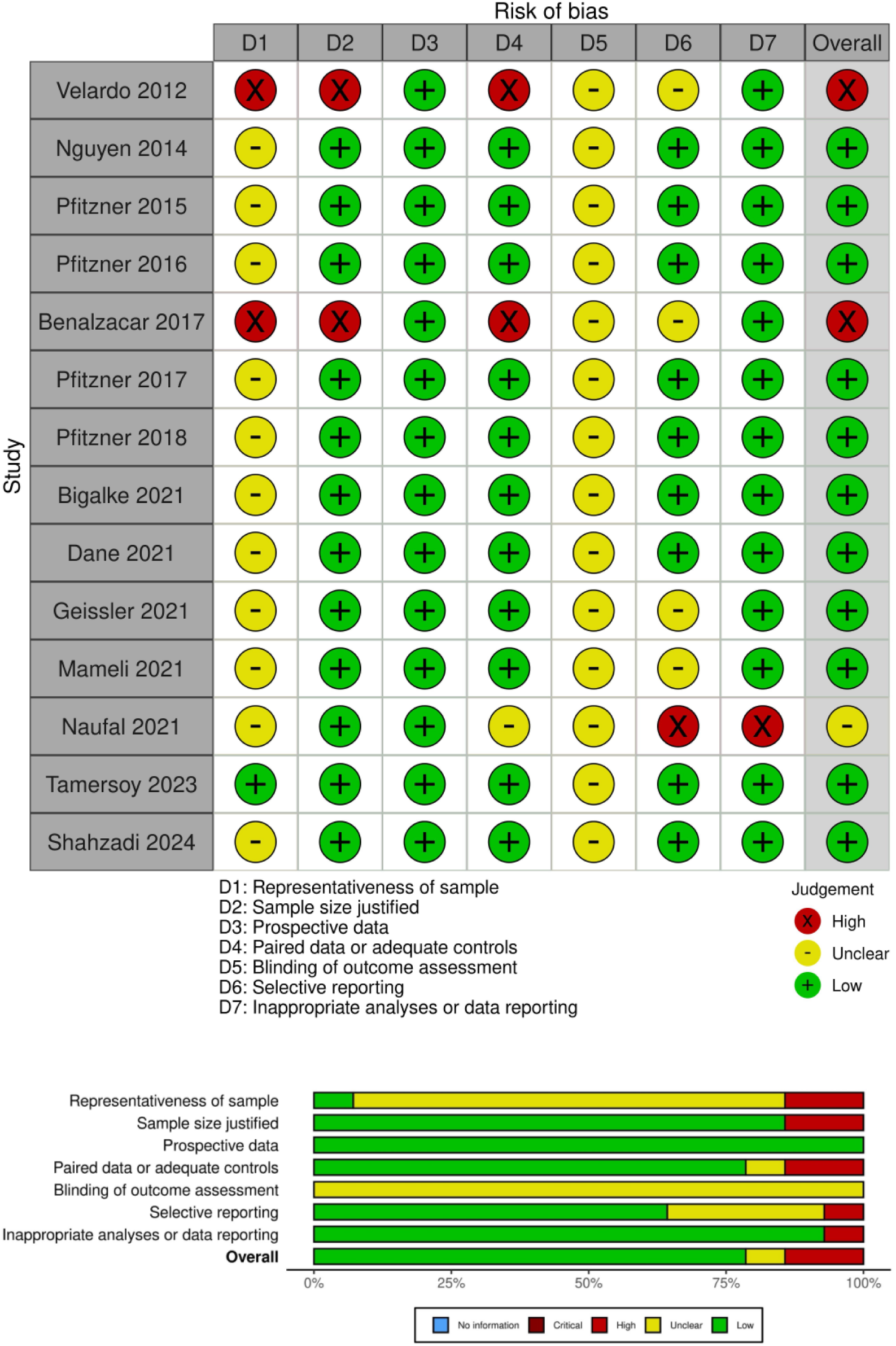
Risk of bias traffic light plot and summary plot based on the Newcastle-Ottawa score. There were no studies with missing information or critical risks.

Eight studies (57%) employed some form of appropriate internal validation of the developed model: split sample analysis in four studies, and cross validation in four studies. No 3D camera weight estimation system had a true external validation process.

### Camera technology and hardware

The original Microsoft Kinect 1 camera was used in 7/14 (50%) studies, and the Microsoft Kinect 2 camera was used in 3/14 (21%) studies. A Siemens FAST 3D camera, a Siemens myExam 3D camera, and an Orbbec Astra camera were used in one study each (7%). The type of 3D camera used was not reported in two studies (14%).

### Fundamental approach used in the weight estimation methodology

Multiple differences approaches were used to process 3D images and obtain a weight estimate from the depth data (see Table 2). Deep learning methods were used in the image preprocessing phase in 4/14 (29%) studies, and in the weight estimation phase in 9/14 (64%) studies. Most methods (9/14 (64%)) required no user input to facilitate the weight estimate calculations, with the exceptions of the methods of Pfitzner and colleagues which required gender as a manual input, and the method of Benalcazar *et al* which required information on clothing and hairstyle.

**Table 2.**
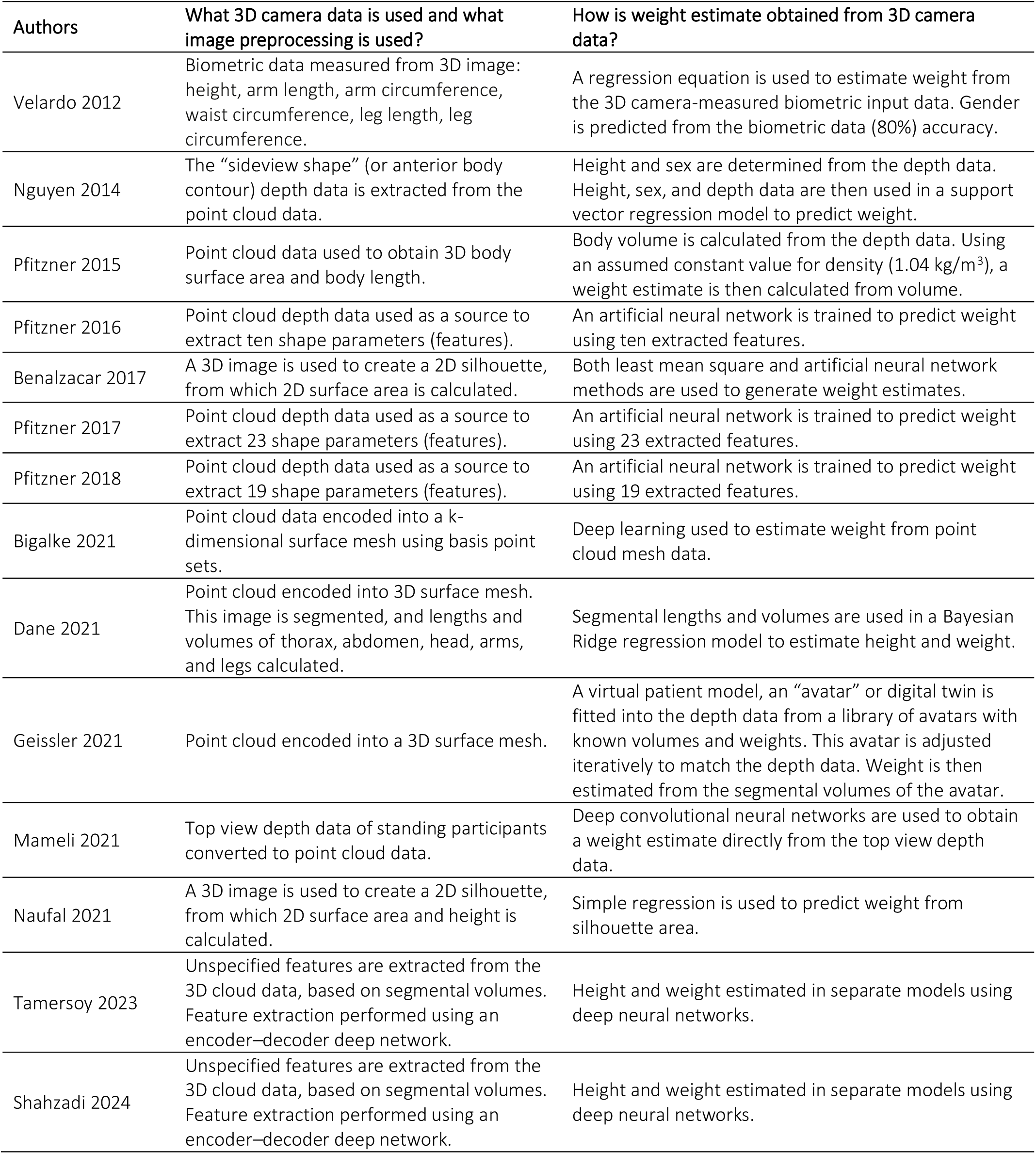
Analytical approach to total body weight estimation in adults.

### Accuracy of weight estimates

Unfortunately, only half of the studies (7/14 (50%)) provided comprehensive data on the performance of the weight estimation systems. Accuracy data (P10 – the percentage of estimates within 10% of actual weight) could be imputed in four additional studies. The accuracy data for each study is shown in Figure 3. Every study for which data was available exceeded the minimum acceptable accuracy standard of P10 >70% [10, 25].

**Figure 3.**
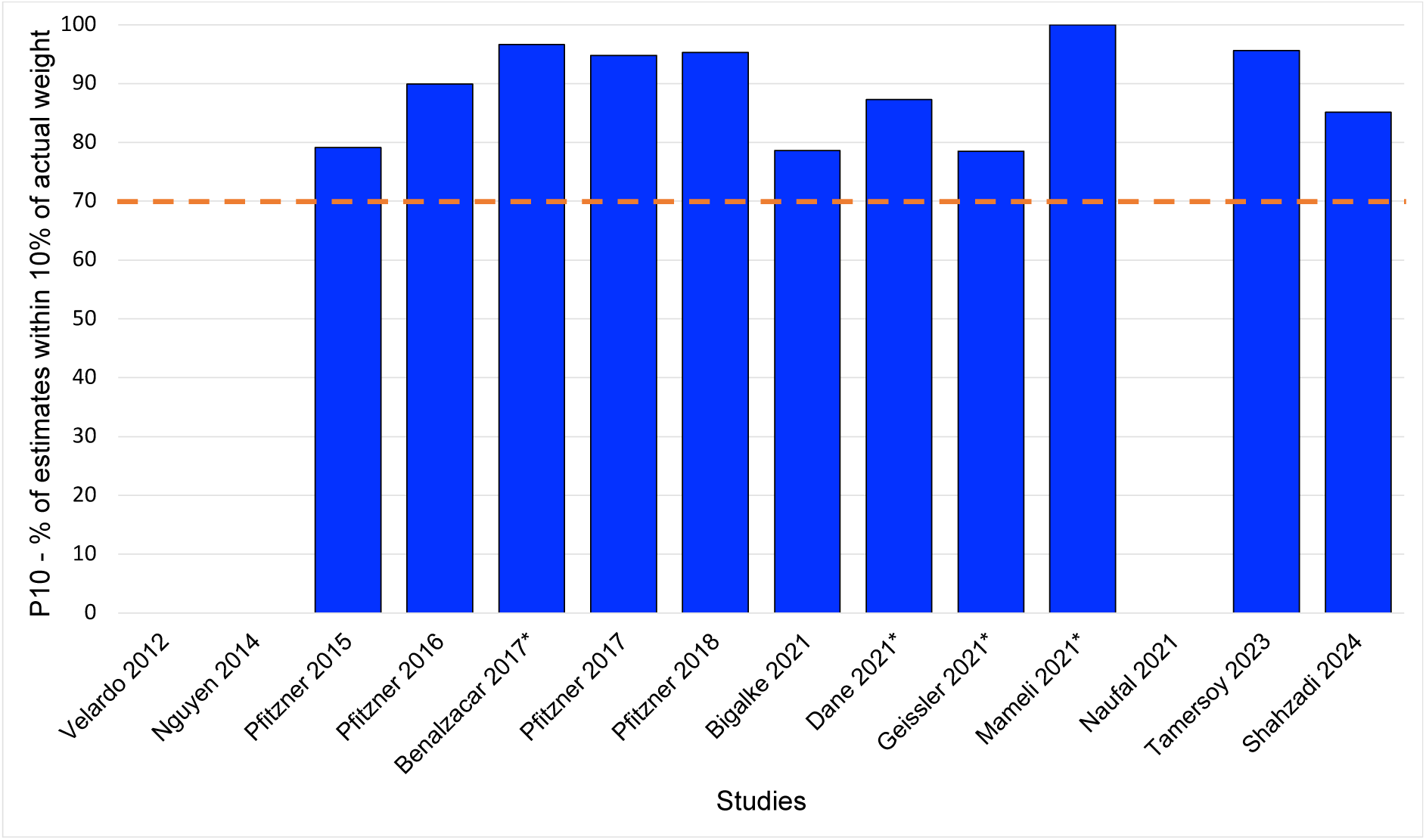
The accuracy data (P10 – percentage of estimates within 10% of actual weight) for each of the 3D camera weight estimation systems. The studies marked with an asterisk (*) identify studies for which P10 data was imputed. The red dashed line indicates the minimum acceptable performance threshold of P10 = 70%.

In the four studies in which direct, paired comparisons were made against other weight estimation systems, the following findings were notable: firstly, the 3D camera systems were always more accurate than guesstimates by healthcare providers (four studies); secondly, the 3D camera systems were always less accurate than participant self-estimates of weight (three studies). Comparative data was not available from the studies in which the 3D cameras achieved exceptionally high accuracy results.

### The suitability of 3D camera weight estimation systems for emergency and critical care

There were 4/14 (29%) studies conducted in an environment designed to simulate an Emergency Department setting, and 5/14 (36%) studies conducted in, or with data from, a radiological suite. However, no study evaluated an estimation method during the provision of actual or simulated emergency care.

## LIMITATIONS

There were several limitations to this review. Firstly, papers in the non-medical literature are less well indexed and searchable than in the medical literature. It is therefore possible that some relevant studies were missed. Secondly, the studies were from a very narrow range of geographical locations, which could limit the generalizability of the findings. Thirdly, the small sample sizes, the variable data reporting and statistical analysis, especially of subgroups of BMI, limited any comparisons between different 3D camera weight estimation systems. The need to impute data was also a limitation. Furthermore, few studies included a sufficiently diverse sample of participants with different ages, ethnic groups, height and weight ranges, and weight status (e.g., underweight, healthy weight, overweight, obese).

## DISCUSSION

The current understanding of 3D camera-based weight estimation in adults, including its potential role during emergency care, has significant gaps. For example, when faced with a critically ill or injured patient in need of urgent weight-based drug therapy, but without any recorded weight, could a 3D camera system be used for estimating their weight? The significance of our review lies in its exploration of the currently available information on this topic. Our aim was to offer information and guidance to clinicians and researchers in this matter of important patient safety. The importance of the topic lies in the imperative for accurate of drug dosing: both treatment failure from underdosing and adverse events from overdosing can be significant threats to life.

We identified and reviewed all the published literature on 3D camera weight estimation methods that could potentially be used during emergency medical care of adult patients. While some methods were primarily intended for nutritional assessment, others were devised and intended to guide acute medical interventions (e.g., to guide dosage of thrombolytic therapy in patients with acute ischemic stroke).

### Quality of the studies

Although a few studies had inadequate data reporting and statistical analysis, most of the studies were methodologically sound. This provided a good evidence basis from which to draw preliminary conclusions. The lack of true external validation studies was a significant limitation in the field of 3D weight estimation, however.

### Camera technology

The studies in this review made use of several different types of 3D cameras: structured light systems (e.g., Microsoft Kinect 1, Orbbec Astra) and time-of-flight systems (e.g., Microsoft Kinect 2). These camera systems are relatively old and, in some cases, no longer manufactured. Newer cameras have native software to perform many image processing tasks automatically: intrinsic and extrinsic calibration to ensure accurate depth measurements and color-depth alignment; automatically correct lens distortion; generate 3D point clouds from depth data; convert depth images to 3D coordinates automatically; detect and track human skeletons in real-time; automatically detect and track objects or faces within the camera’s field of view; provide bounding boxes or other positional data for detected objects; apply noise reduction and smoothing filters to depth data; perform edge detection and other image processing tasks. These newer 3D cameras are, therefore, likely to be better than those already tested. Existing research relating to the cameras themselves has been sparse, and future work needs to evaluate the most appropriate hardware system for use for weight estimation and in potential clinical emergency medicine applications. Table 3 provides a description of the different types of 3D cameras.

**Table 3.**
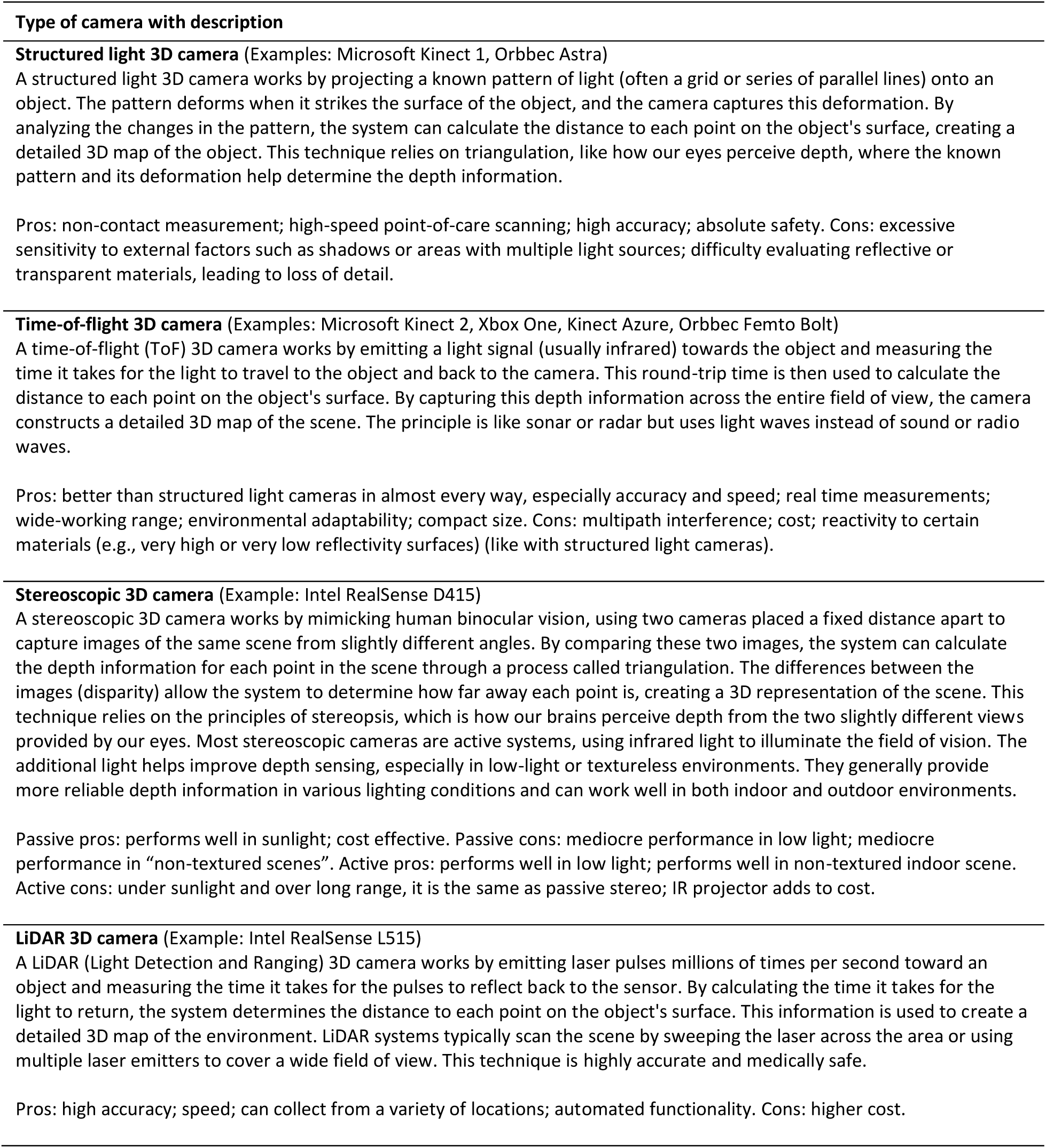
3D cameras used in the weight estimation systems.

| Type of camera with description |
| --- |
| **Structured light 3D camera** (Examples: Microsoft Kinect 1, Orbbec Astra)<br>A structured light 3D camera works by projecting a known pattern of light (often a grid or series of parallel lines) onto an object. The pattern deforms when it strikes the surface of the object, and the camera captures this deformation. By analyzing the changes in the pattern, the system can calculate the distance to each point on the object's surface, creating a detailed 3D map of the object. This technique relies on triangulation, like how our eyes perceive depth, where the known pattern and its deformation help determine the depth information.<br>Pros: non-contact measurement; high-speed point-of-care scanning; high accuracy; absolute safety. Cons: excessive sensitivity to external factors such as shadows or areas with multiple light sources; difficulty evaluating reflective or transparent materials, leading to loss of detail. |
| **Time-of-flight 3D camera** (Examples: Microsoft Kinect 2, Xbox One, Kinect Azure, Orbbec Femto Bolt)<br>A time-of-flight (ToF) 3D camera works by emitting a light signal (usually infrared) towards the object and measuring the time it takes for the light to travel to the object and back to the camera. This round-trip time is then used to calculate the distance to each point on the object's surface. By capturing this depth information across the entire field of view, the camera constructs a detailed 3D map of the scene. The principle is like sonar or radar but uses light waves instead of sound or radio waves.<br>Pros: better than structured light cameras in almost every way, especially accuracy and speed; real time measurements; wide-working range; environmental adaptability; compact size. Cons: multipath interference; cost; reactivity to certain materials (e.g., very high or very low reflectivity surfaces) (like with structured light cameras). |
| **Stereoscopic 3D camera** (Example: Intel RealSense D415)<br>A stereoscopic 3D camera works by mimicking human binocular vision, using two cameras placed a fixed distance apart to capture images of the same scene from slightly different angles. By comparing these two images, the system can calculate the depth information for each point in the scene through a process called triangulation. The differences between the images (disparity) allow the system to determine how far away each point is, creating a 3D representation of the scene. This technique relies on the principles of stereopsis, which is how our brains perceive depth from the two slightly different views provided by our eyes. Most stereoscopic cameras are active systems, using infrared light to illuminate the field of vision. The additional light helps improve depth sensing, especially in low-light or textureless environments. They generally provide more reliable depth information in various lighting conditions and can work well in both indoor and outdoor environments.<br>Passive pros: performs well in sunlight; cost effective. Passive cons: mediocre performance in low light; mediocre performance in "non-textured scenes". Active pros: performs well in low light; performs well in non-textured indoor scene. Active cons: under sunlight and over long range, it is the same as passive stereo; IR projector adds to cost. |
| **LiDAR 3D camera** (Example: Intel RealSense L515)<br>A LiDAR (Light Detection and Ranging) 3D camera works by emitting laser pulses millions of times per second toward an object and measuring the time it takes for the pulses to reflect back to the sensor. By calculating the time it takes for the light to return, the system determines the distance to each point on the object's surface. This information is used to create a detailed 3D map of the environment. LiDAR systems typically scan the scene by sweeping the laser across the area or using multiple laser emitters to cover a wide field of view. This technique is highly accurate and medically safe.<br>Pros: high accuracy; speed; can collect from a variety of locations; automated functionality. Cons: higher cost. |

### Analytical approach to weight estimation

The analytical approaches to weight estimation have evolved significantly in successive studies over the last decade. The earliest system described the use of a 3D camera to obtain biometric data which could be used in an equation derived from an anthropometric dataset [11]. Subsequent studies used depth data to calculate total body volumes (and, later, segmental body volumes), which are converted to weight estimates using density constants [13, 14, 16, 17]. The most recent methodologies have used deep learning to match a digital twin from a library of trained images against the point cloud data of a captured 3D image [24, 26]. This is perhaps the most flexible method, with the highest potential for accuracy. The use of deep learning both in image processing and in the weight estimation process has substantially improved the accuracy of weight estimates.

### Accuracy of weight estimation by 3D camera systems

The best metric for evaluating the global performance of a weight estimation system is the overall accuracy (P10 and/or P20) [10]. In this review, each of the 11 weight estimation systems for which P10 data was available exceeded the minimum required accuracy threshold for a weight estimation system (P10>70%), as has been described previously [9, 25]. In fact, the lowest P10 was just below 80%, and four systems had a P10>95%. Overall, this performance data is remarkably good. To put this in context, a recent meta-analysis of weight estimation systems in adults showed that only patient self-estimates of weight approached this degree of accuracy but were inconsistent across studies [4]. In addition, self-estimates of weight were often not able to be provided by the sickest patients. In studies conducted during actual emergency medical situations, the number of patients unable to provide a self-estimate may be as high as 70 to 85% [27]. The evidence is thus clear that methods of weight estimation that do not rely on self-estimates must always be available [4]. The data from this scoping review shows that 3D camera systems could potentially fulfil this role if their performance holds up in larger scale clinical studies. The limited subgroup data presented suggested that weight estimation accuracy may be maintained in patients with obesity, but that models might need to improve their performance in patients who are underweight.

Other noteworthy factors were that accurate weight estimation was achieved with several different 3D cameras, as well as with different processing and analytical approaches. This strongly supports the validity of the underlying principles, and predictable biological associations between body size, shape, and body weight. In addition, accurate weight estimation was even possible when patients were clothed or covered with light blankets [28].

### Appropriateness for use during ED or prehospital emergency care

The appropriateness of 3D camera weight estimation systems for use during emergency care was not explicitly studied, although several of the studies specifically intended their systems to be used for this purpose [13, 14, 16, 17, 29]. There are several factors that make fully evolved 3D camera systems ideal for use during emergency medical care. Firstly, they are quick. A weight estimation can be calculated in less than one second, even with the use of deep learning systems in both the image preprocessing and the weight estimation algorithms [16]. Secondly, they are highly automated. The system can automatically select the optimum image to use for the processing (useful for when patients are moving or uncooperative). No user input is required for the weight estimation: sex and height, which have significant associations with weight, can be estimated using deep learning. Thirdly, the system can compensate for patient posture and patient movement. Irrespective of whether the patient is supine, prone, or lateral, an accurate weight estimate can be obtained. Finally, light clothing or coverings do not interfere with weight estimation, as 3D camera systems can “see beneath the covers” using deep learning digital twin-based analyses.

### Future directions

This is an important and exciting field for future research. The 3D camera systems need to be studied in larger samples, including representative numbers of underweight and obese patients, as well as patients from diverse population groups, to ensure generalizability. These methodologies also need to be evaluated in clinical environments, including during emergency care. Likewise, the research needs to include children. Future innovations could also include the estimation of ideal body weight and lean body weight to allow for precision weight-based dosing for all patients. At present, establishing these weights is complex and requires additional measurements and calculations. They are thus not routinely employed by emergency physicians.

## CONCLUSIONS

The weight estimation accuracy of 3D camera-based systems represents a significant advancement in the field of automated measurement and analysis. These systems utilize precise depth sensing and 3D modeling to capture the volume and dimensions of objects or individuals with high accuracy. By integrating advanced algorithms and machine learning techniques, 3D camera-based systems can convert depth data into reliable weight estimates. When properly optimized, 3D camera-based weight estimation can achieve accuracy comparable to traditional weighing methods, providing a non-contact, efficient, and versatile potential solution for use during emergency care. However, it was clear from this review that additional, high quality prospective research is urgently needed in this field, as a matter of prioritizing patient safety during emergency care.

## Data Availability

All data produced in the present study are available upon reasonable request to the authors

## Notes

### Competing Interest Statement

The authors have declared no competing interest.

### Funding Statement

This study did not receive any funding

### Summary of Updates

The author list was updated with the correct version..

