## Supplementary Table 1 for "Total body weight estimation by 3D camera systems: potential high-tech solutions for emergency medicine applications? A scoping review" (from another version of this preprint: version 1, 2024-08-15)

Supplementary Table 1 Details of the search strategy.

| | Search Terms |
|---|---|
| **Potential patients (population)*** | 1. adult/ OR young adult/ OR adolescent/ OR child/ OR infant/ MeSH<br>2. (human) [Title/Abstract]<br>3. (livestock OR pig OR cattle OR cow OR sheep OR lamb OR horse OR fish OR vegetable OR lettuce OR pumpkin OR fetus OR fetal OR kidney OR renal OR ventricular) [Title/Abstract].<br>4. 1 OR 2 **NOT** 3 |
| **3D camera (exposure)** | 5. (3D OR 3-D) [Title/Abstract].<br>6. ("3-dimensional" OR "three dimensional" OR "three-dimensional") [Title/Abstract].<br>7. (RGBD OR RGB-D OR (RGB)-D OR IR-UWB) [Title/Abstract].<br>8. ("depth camera" OR "depth-camera" OR Kinect OR "computer vision") [Title/Abstract].<br>9. 5 or 6 or 7 or 8 |
| **Weight estimation (outcome)** | 10. body weight/ OR ideal body weight/ MeSH<br>11. ("weight estimation" OR "estimating weight" OR "weight prediction" OR "estimation of weight" OR "estimating body weight" OR "weight measurement") [Title/Abstract].<br>12. (silhouette OR "patient avatar" OR "digital twin") [Title/Abstract].<br>13. 10 OR 11 OR 12 OR 13 |
| **Combined** | 14. 4 AND 9 AND 13 |
| MEDLINE: (((("3D"[Title/Abstract] OR "3-D"[Title/Abstract] OR "3-dimensional"[Title/Abstract] OR "three-dimensional"[Title/Abstract] OR "three-dimensional"[Title/Abstract] OR "RGBD"[Title/Abstract] OR "RGB-D"[Title/Abstract] OR "RGB"[All Fields]) AND "d"[Title/Abstract]) OR "IR-UWB"[Title/Abstract] OR "depth-camera"[Title/Abstract] OR "depth-camera"[Title/Abstract] OR "Kinect"[Title/Abstract] OR "computer vision"[Title/Abstract]) AND ("body weight"[MeSH Terms] OR "ideal body weight"[MeSH Terms] OR ("weight estimation"[Title/Abstract] OR "estimating weight"[Title/Abstract] OR "weight prediction"[Title/Abstract] OR "estimation of weight"[Title/Abstract] OR "estimating body weight"[Title/Abstract] OR "weight measurement"[Title/Abstract] OR "silhouette"[Title/Abstract] OR "patient avatar"[Title/Abstract] OR "digital twin"[Title/Abstract]))) NOT ("pig"[Title/Abstract] OR "cattle"[Title/Abstract] OR "cow"[Title/Abstract] OR "sheep"[Title/Abstract] OR "lamb"[Title/Abstract] OR "horse"[Title/Abstract] OR "fish"[Title/Abstract] OR "vegetable"[Title/Abstract] OR "lettuce"[Title/Abstract] OR "pumpkin"[Title/Abstract] OR "fetus"[Title/Abstract] OR "fetal"[Title/Abstract] OR "kidney"[Title/Abstract] OR "renal"[Title/Abstract] OR "ventricular"[Title/Abstract]) | |
| Embase: ("weight estimation" OR "estimating weight" OR "weight prediction" OR "estimation of weight" OR "estimating body weight" OR "silhouette" OR "patient avatar") AND (3D OR 3-D OR "depth camera" OR Kinect) | |
| IEEE: ("weight estimation" OR "estimating weight" OR "weight prediction" OR "estimation of weight" OR "estimating body weight" OR "silhouette" OR "patient avatar") AND (3D OR 3-D OR "depth camera" OR Kinect) | |
| Google: (weight AROUND (3) estimate OR weight AROUND (3) estimation) AND ("3D camera" OR "depth camera" OR "RBGD camera" OR "computer vision")-pig-lamb-fish-livestock-cattle | |
