## Supplementary Table 2 for "Total body weight estimation by 3D camera systems: potential high-tech solutions for emergency medicine applications? A scoping review" (from another version of this preprint: version 1, 2024-08-15)

Supplementary Table 2 Newcastle-Ottawa Scale adapted for weight estimation accuracy studies. Each study is rated in three major categories and can achieve a maximum of 10 stars overall. The number of stars represents the methodological strength of the study.

Selection: (Maximum 4 stars)

1. Representativeness of the study sample:
    a. Truly representative of the target population (all subjects or random sampling with large sample size) (★★)
    b. Somewhat representative of the target population (non-random sampling or small consecutive sampling or relevant convenience sampling) (★)
    c. Selected group or no description of the sampling strategy.
2. Sample size:
    a. Justified and acceptable (★)
    b. Not justified or not acceptable.
3. Prospective data (★)

Comparability of cohorts or study design or analysis: (Maximum 2 stars)

1. Paired data or appropriate evaluation of single weight estimation method (★★)
2. Study controls adequately for additional factors (★)

Outcome: (Maximum 4 stars)

1. Assessment of the outcome:
    a. Independent blinded assessment (★★)
    b. Independent non-blinded assessment (★)
    c. Non-blinded assessment or no description of assessment

If data analysis was not appropriate for method-comparison studies (no assessment of bias, precision (or repeatability if applicable) and accuracy) then consider downgrading one level.

2. Statistical test:
    a. Appropriate, clearly described statistical analysis with confidence intervals or measure of variance, with the use of percentage error or logarithmic transformation analysis and with appropriate use of relevant and important subgroup analyses (★★)
    b. Appropriate, clearly described statistical analysis with confidence intervals or measure of variance (★)
    c. The statistical testing is incomplete.
    d. The statistical testing is inappropriate, does not support conclusions (-1★)
